# Exploring *Blastocystis* and Gut Health: A Pilot Investigation of Microbiome and Metabolomic Signatures in a UK Cohort

**DOI:** 10.1101/2025.08.19.25333969

**Authors:** William JS. Edwards, Jamie M. Newton, Gary Thompson, Eleni Gentekaki, Anastasios D. Tsaousis

## Abstract

*Blastocystis* is a common protistan coloniser of the human gut, yet its role in health and disease remains unclear. While some studies have linked it to gastrointestinal (GI) symptoms, growing evidence suggests an association with microbial diversity and gut health. This pilot study investigated the prevalence, subtype distribution, and ecological associations of *Blastocystis* in South East England, and explored its relationship with gut microbiota composition and metabolic profiles, particularly in individuals with or without irritable bowel syndrome (IBS) or ulcerative colitis (UC). We analysed stool samples from 46 participants using qPCR and sequencing to identify *Blastocystis* subtypes, 16S rRNA gene sequencing to characterise bacterial communities, and ^1^H NMR spectroscopy to profile the faecal metabolome. *Blastocystis* was detected in 47.8% of individuals, with subtypes ST1–ST4 being the most frequently identified. Alpha diversity was higher in *Blastocystis*-positive individuals, significantly so in IBS cases. While *Blastocystis* status did not alter overall microbial or metabolomic beta diversity substantially, disease status (IBS/UC) did. Notably, several *Blastocystis*-associated taxa were negatively correlated with leucine and kynurenine metabolites implicated in inflammation and GI dysfunction. Leucine was positively associated with IBS status, while kynurenine is a known pro-inflammatory product of tryptophan metabolism. These correlations suggest that *Blastocystis* colonisation may reflect or contribute to gut environments with lower inflammatory potential. This study highlights the need for larger, longitudinal and subtype-resolved investigations to clarify the ecological and functional roles of *Blastocystis* in the gut microbiome. Our findings support the growing view that *Blastocystis*, particularly certain subtypes, may be more indicative of gut health than disease.

## Introduction

*Blastocystis* is one of the most commonly detected microbial eukaryotes in the human gut, colonising over a billion people worldwide (Stensvold, 2012). It has adapted to a gut lifestyle and has a broad host range, infecting not only humans but also a variety of animal species ((Eme *et al*., 2017)(Alfellani *et al*., 2013)(Udonsom *et al*., 2018)(Betts *et al*., 2018)). Despite its broad distribution, the clinical relevance of *Blastocystis* remains unclear (Tsaousis, Gentekaki and Stensvold, 2024)(Stensvold, 2025). The organism was historically classified as a pathogen, and links to gastrointestinal (GI) symptoms and disorders such as irritable bowel syndrome (IBS) and inflammatory bowel disease (IBD) were proposed, but later challenged (Poirier *et al*., 2012)(Krogsgaard *et al*., 2015). Nonetheless, recent studies on background populations have revealed that the organism occurs with high frequency in healthy individuals without GI symptoms shifting the view towards *Blastocystis* being a part of the gut healthy microbiome (Andersen *et al*., 2015)(Jinatham *et al*., 2021)(McCain *et al*., 2023)(Stensvold and van der Giezen, 2018).

*Blastocystis* possesses extreme genetic diversity (Denoeud *et al*., 2011)(Gentekaki *et al*., 2017)(Stensvold *et al*., 2023). At least 45 subtypes (STs) have been described to date, with differences in host range (Stensvold and Clark, 2020; Baek et al., 2022). Of these, ST1-ST4 are the typical subtypes of humans though others have also been sporadically detected (Rauff-Adedotun *et al*., 2021)(Nguyen *et al*., 2023).

In the past decade, several studies associated *Blastocystis* colonisation with a more diverse and compositionally richer gut microbiome in gut healthy individuals (Nieves-Ramírez *et al*., 2018)(Audebert *et al*., 2016)(Kodio *et al*., 2019). More recently, potential links between Blastocystis subtypes, microbiota profiles and population parameters were explored.

The role of *Blastocystis* and the various subtypes in the gut is being studied intensively, though most studies remain observational (Beghini *et al*., 2017)(Tito *et al*., 2019)(Piperni *et al*., 2024). For instance, several studies (Tito *et al*., 2019)(Yason *et al*., 2019)found that individuals harbouring *Blastocystis*—particularly ST4—exhibited higher microbial richness and abundance of commensal taxa compared to those without. These findings have now been reinforced by a landmark global metagenomic study (Piperni *et al*., 2024). Importantly, its presence was strongly associated with increased bacterial alpha diversity, enrichment of health-associated taxa, and a reduction in dysbiosis-associated signatures. The organism was more common in individuals with high-fibre diets, lower inflammation markers, and overall gut microbiome configurations consistent with health. These findings strongly support the hypothesis that *Blastocystis*, particularly specific subtypes, may serve as a sentinel of gut ecosystem stability and eubiosis.

In addition to microbial composition, the gut metabolome offers insights into the functional consequences of *Blastocystis* colonisation. Metabolites produced through host–microbiota interactions play key roles in immune signalling, nutrient processing, and gut barrier integrity. Metabolomic profiling may therefore provide valuable information about how *Blastocystis* influences (or is influenced by) gut ecosystem function. Some studies have shown that *Blastocystis* colonisation can modulate the abundance of key (Betts *et al*., 2021), though the mechanisms and clinical significance of these interactions remain largely unexplored.

Given the increasing evidence linking *Blastocystis* with both health and disease, and the likely subtype-specific effects, population-based studies are essential. South East England represents a region with limited epidemiological data on *Blastocystis* prevalence, subtype distribution, and associations with GI health. In this pilot study, we aimed to: (1) determine the occurrence and subtype distribution of *Blastocystis* in a mixed cohort of individuals with and without IBS in South East England; (2) assess its association with gut bacterial diversity and composition using 16S rRNA gene amplicon sequencing; and (3) examine its relationship with gut metabolite profiles using ^1^H NMR spectroscopy. By integrating microbiome and metabolome data, this study seeks to provide preliminary insights into the ecological role of *Blastocystis* and its relevance to gut health in a UK-based population.

## Methods

### Ethics Statement and research permissions

The study was conducted within the guidelines established in IRAS ethics approvals 274985 and 286641, following a review by an ethics committee and applying suggested amendments to comply with ethical standards. Final approval was granted by the Health Research Authority (HRA) following a review of the project and the application of amendments suggested by the HRA to comply with their ethical standards.

### Study area and sample collection

All participants were recruited from regions within the U.K., with all but two residing in Kent (UK). Participants were recruited via three channels. IBS participants were recruited through Crohn’s and Colitis UK (https://crohnsandcolitis.org.uk/our-work/research-andevidence), gastroenterologists at William Harvey Hospital (Ashford, Kent, UK) and advertisements in supermarkets, doctors’ surgeries, and other community-targeted channels. Non-IBS participants were recruited using the same community-targeted channels as IBS participants. Participants were provided with faeces catchers and two faeces collection tubes for each collection, one containing 5 mL of DNA/RNA shield and the other containing 5 mL of 50% methanol. Faecal samples were collected at home by the participants, then transported to the lab via mail, courier or delivered in person, and then stored at -80°C in their respective tubes in 50% methanol or DNA/RNA shield (Cambridge Biosciences). All participants were sent a questionnaire in which information about a set of demographic, health and lifestyle factors was acquired.

### Genomic DNA Extraction

The samples stored in DNA/RNA shield were thawed, then 200 mg solid stool or 200 µl liquid stool were added to 200 µL PBS (pH 7.4 RNAase free). The samples were then centrifuged for 10 minutes at 10,000 g at room temperature (RT). The pellet was then resuspended in the supernatant, and the DNA was extracted using the QIAamp PowerFecal Pro DNA Kit (Qiagen, Catalogue no 51804) following the manufacturer’s protocol, and 50 µL of DNA was eluted. DNA concentrations were measured by nanodrop using 2 µL DNA.

### qPCR (Real-time PCR)

For qPCR *Blastocystis* detection a 350 bp barcoding region of the SSU rRNA gene was targeted using a reaction mixture of 2 µL DNA, 500 nM of primer set PPF1 (fwd) (5’-AGTAGTCATACGCTCGTCTCAAA-3’) and R2PP (rvs) (5’-TCTTCGTTACCCGTTACTGC-3’) and 5 µL SYBR green making a full reaction volume of 10 µL. The qPCR was run on a QuantStudio 3 real-time PCR machine with the following program; Initial denaturation 95 °C for 5 minutes, then 45 cycles of initial denaturation 95°C 5 seconds, annealing 68 °C 10 seconds, extension 72 °C 10 seconds then a final extension of 72 °C 15 seconds.

### Barcoding Nested PCR

For nested PCR a barcoding region of the SSU rRNA gene was also targeted by using two consecutive PCR cycles. The conditions for the first cycle were as follows: 2 µL extracted DNA for Blastocystis detection, 10 µL 5 x buffer (Promega), 1 mM MgCl2, 0.4 µM of primer set RD3 (fwd) 5′-GGGATCCTGATCCTTCCGCAGGTTCACCTAC-3′ and RD5 (rvs) 5′-GGAAGCTTATCTGGTTGATCCTGCCAGTA-3′ (Clark, 1997), 0.2 mM dNTPs (Promega), 0.25 µL Taq polymerase, 30.75 µL sterile H2O comprising a 50 µL reaction. The second nested PCR was performed under the same conditions as above with 1 µL extracted DNA and 31.75 µL sterile H2O using the primer set RD5F (fwd) 5′-ATCTGGTTGATCCTGCCAGT-3′ and BhRDr (rvs) 5′-GAGCTTTTTAACTGCAACAACG-3′, producing a fragment of approximately 650 bp, considered the barcoding region. Both PCRs were run under the following cycling conditions: Initial denaturation at 95 °C for 5 minutes, then 35 cycles of denaturation at 95 °C for 30 s, annealing at 55 °C for 30 s, extension at 72 °C for 1 minute and 40 s and final extension at 72 °C minute.

### Gel extraction, Cloning and sequencing

The positive qPCR reactions were purified using the Qiagen QIAquick PCR Purification Kit (Qiagen) following the manufacturer’s protocol. The positive nested PCR reactions were gel extracted using the Qiagen QIAquick Gel Extraction Kit following the manufacturer’s protocol. The nested PCR reactions were cloned into a pGEM-T vector (Promega) following the manufacturer’s protocol. One to seven colonies were selected from each transformation and subcultured. Then the plasmid was purified using the Qiagen QIAprep Spin Miniprep Kit. Purified plasmids were then screened to confirm the presence of the 650 bp fragment by digestion with the restriction enzyme *EcoRI*. Bidirectional Sanger sequencing for the qPCR reactions, gel extractions and positive plasmids using the primer sets PPF1 and R2PP, RD3 and BhRDr and RD3 and BhRDr, respectively, was outsourced to Eurofins UK. The forward and reverse nucleotide sequences were then assessed and trimmed if necessary with, SnapGene Viewer Version 6.2.2 (https://www.snapgene.com/snapgene-viewer). The final trimmed consensus sequence was then compared to reference sequences from GenBank using the Basic Local Alignment Search Tool (BLAST) from the National Center for Biotechnology Information (NCBI) (https://blast.ncbi.nlm.nih.gov/Blast.cgi). The subtypes were then confirmed using the *Blastocystis* database at https://pubmlst.org/multilocus-sequence-typing.

### 16S rRNA Gene Amplicon Sequencing

High-throughput amplicon sequencing was performed by Novogene, utilising the methodology of Caporaso et al. (Caporaso *et al*., 2011). DNA was fragmented then adapters added for paired-end sequencing. The target gene was the V3-V4 region of the 16S rRNA gene amplified using the 515F (GTGCCAGCMGCCGCGGTAA) and 907R (CCGTCAATTCCTTTGAGTTT) primer pair.. Sequencing was performed using the Illumina NovaSeq platform.

Raw 16s reads were processed into ASVs using the Lotus2 pipeline (Özkurt *et al*., 2022). Sequences underwent Chimera detection/removal and identification and exclusion of off-target human DNA contaminated reads (BLAST search against the Genome Reference Consortium Human Build 38.p14), was performed using Minimap2 (Li, 2018). The V3-V4 region of the trimmed reads was then clustered into Amplicon Sequence Variants (ASVs), using the Divisive Amplicon Denoising Algorithm 2 (DADA2) (Callahan *et al*., 2016). ASVs classified to the species level via BLAST against the GreenGenes2 (GG2) database (DeSantis *et al*., 2006). The database was chosen due to its status as a unified database suitable for both whole-genome sequencing (WGS) and 16S rRNA sequencing, as well as the thorough chimaera checking of its sequences, which ensures reproducibility of results.

### Statistical Analysis

Statistical analyses of the 16S sequences (as ASVs) and data visualisation were performed using R Studio 4.2.3. due to varying sequencing depths across samples, sample data was rarefied (60,000 reads). Diversity indices,; Shannon, Chao1, Simpson, and observed taxa, were computed using the Phyloseq package and were used to assess differences between *Blastocystis*-positive and *Blastocystis*-negative samples. Statistical testing on the diversity indices was done with either one-way ANOVA, followed by the Tukey HSD test (for normally distributed data) or the Kruskal-Wallis test followed by the Dunn test (with Bonferroni adjustment) (for non-normally distributed data). Compositional bar plots were visualised using the Microbiome package, incorporating only taxa representing more than 1% of the total read count within each sample. Principal Coordinate Analysis (PCoA) was then performed to assess differences in microbial community structure between *Blastocystis*-positive and *Blastocystis*-negative samples utilising non-Euclidean distance matrices. Samples in accordance with their Bray-Curtis dissimilarity matrices, and PERMANOVA (Anderson, 2017) was used to statistically confirm if centroids of the Blastocystis status groups significantly differed in location. To investigate potential biomarkers, LEfSe (Segata *et al*., 2011) was used to identify linear discriminant taxa. LEfSe was used as it factors in both abundance and prevalence and accounts for multiple testing. The results of LEfSe were then correlated with the differentially abundant metabolites identified using a Spearman’s correlation plot.

### Metabolite extraction

200 mg of solid stool was taken from the stool samples suspended in 50% methanol and resuspended in a fresh tube of 4 ml of 50% methanol containing 200 mg of 2 mm diameter glass beads. The sample was vortexed for 30 s, then incubated at room temperature (RT) for 3 minutes, then vortexed for a further 30 s. The homogenised lysate was separated into 4 x 1 ml aliquots and centrifuged at 10,000 x g for 20 minutes at 4 °C. The supernatants were then transferred into fresh tubes and lyophilised.

### Preparation for 1H NMR acquisition

The lyophilised desiccates were suspended in 330 µL MiliQ H2O then vortexed for 30 s. The four supernatants of each sample were recombined, and 147 µL 10 mM non-deuterated DSS dissolved in D2O was added, resulting in a solution of 10% D2O, 1 mM DSS.

### Metabolite detection from extracts by 1H NMR spectroscopy

A 1H NOESY experiment with a 100 ms mixing time was run on a 600 MHz Avance III NMR spectrometer (Bruker) with a QCI-P cryoprobe to acquire a 1D NMR Spectrum. Experiments were conducted at a calibrated temperature of 298 K. The experiments were run using IconNMR with an automated set of macros. An excitation sculpting experiment was performed for calibration on each sample which included locking to D2O, tuning and matching, measurement of water offset and 90° pulse calibration. The soft pulse power levels were calculated from attenuated values from the 90° pulse. The receiver gain was measured for each sample and limited to 128. The data was acquired over 512 scans with eight dummy scans. A spectral width of 12.02 ppm (7211 Hz) was used and 32,768 data points acquired, giving an acquisition time of 1.27 s, separated by a relaxation delay of 3 s. The acquisition time and relaxation delay were set to give adequate water suppression.

### Processing of ^1^H NMR data

All spectra were phased, baseline corrected, and a 1 Hz window function was applied using TopSpin 3.6.1 (Bruker) software. The spectra were then imported into Chenomx 8.4 and the region between 4.56 ppm and 4.97 ppm was deleted to eliminate the peaks impacted by water resonance. Peaks were assigned using a profiler tool to fit them into the Chenomx library of 338 metabolites. Metabolite concentrations were calculated automatically in Chenomx 8.4 as a proportion of the DSS-d6 standard then fit accurately when the metabolites were assigned. The data comprising of the detected metabolites and their concentrations was then exported from Chenomx 8.4]

### Metabolite analysis

Metabolite data were processed using [INPUT], and chemical concentrations (ppm) were identified. Metabolite data was then visualised using R Studio 4.2.3. The metabolite composition of the samples was first visualised using Principle Component Analysis (PCA) and PERMANOVA (Anderson, 2017) was used to statistically confirm if centroids of the Blastocystis status groups significantly differed in location.

Differentially abundant metabolites were then identified using volcano plots, where log fold change values were plotted against log p-values. Metabolites significantly up- or downregulated in *Blastocystis* +ve/-ve samples were identified and recorded.

## Results

### General demographics of participants

A cohort of 46 participants was studied, and all were sent a questionnaire to acquire demographic information (**Table 1**). Nineteen (46.34%) males and 22 (53.66%) females responded, while five participants did not respond. The five participants who did not respond were still tested for *Blastocystis* presence, subtype, faecal bacterial and faecal metabolome composition. Two participants responded to the questionnaire but refused to give their BMI and daily defecation rate. Age groups were defined in ten-year increments, ranging from 21-30 to over 80, as there were no volunteers in the 18-20 age group. The distribution of age groups in numbers was five (12.2 %), seven (17.1 %), six (14.6 %), nine (22 %), eight (19.6 %), five (12.206 %) and one (2.4%) in age groups 21-30, 31-40, 41-50, 51-60, 61-70, 71 -80 and over 80, respectively. All participants except one lived in the South East of England, 21/46 (45.7 %) had IBS, 13/46 (28.3%) had ulcerative colitis (UC), and 18/46 (39.136 %) owned pets.

### Prevalence and subtype distribution of *Blastocystis*

*Blastocystis* was detected in 22/46 (47.8%) individuals. Of these, two were positive for IBS and for *Blastocystis*, six for IBS, UC and *Blastocystis*, and fourteen were positive for *Blastocystis* but had no IBS or UC. No individuals were positive for UC and *Blastocystis*. A total of six subtypes were detected, namely ST1, ST2, ST3, ST4, ST6, and ST7 (**Figure 1**). The most common subtype was ST3 ((6/22), 27.3%), followed by ST1 ((5/22) 22.7%) and ST2 ((4/22) 18.2%), then ST4 ((3/22) 13.6%) and ST6 and ST7 which each were detected only once.

**Figure 1:**
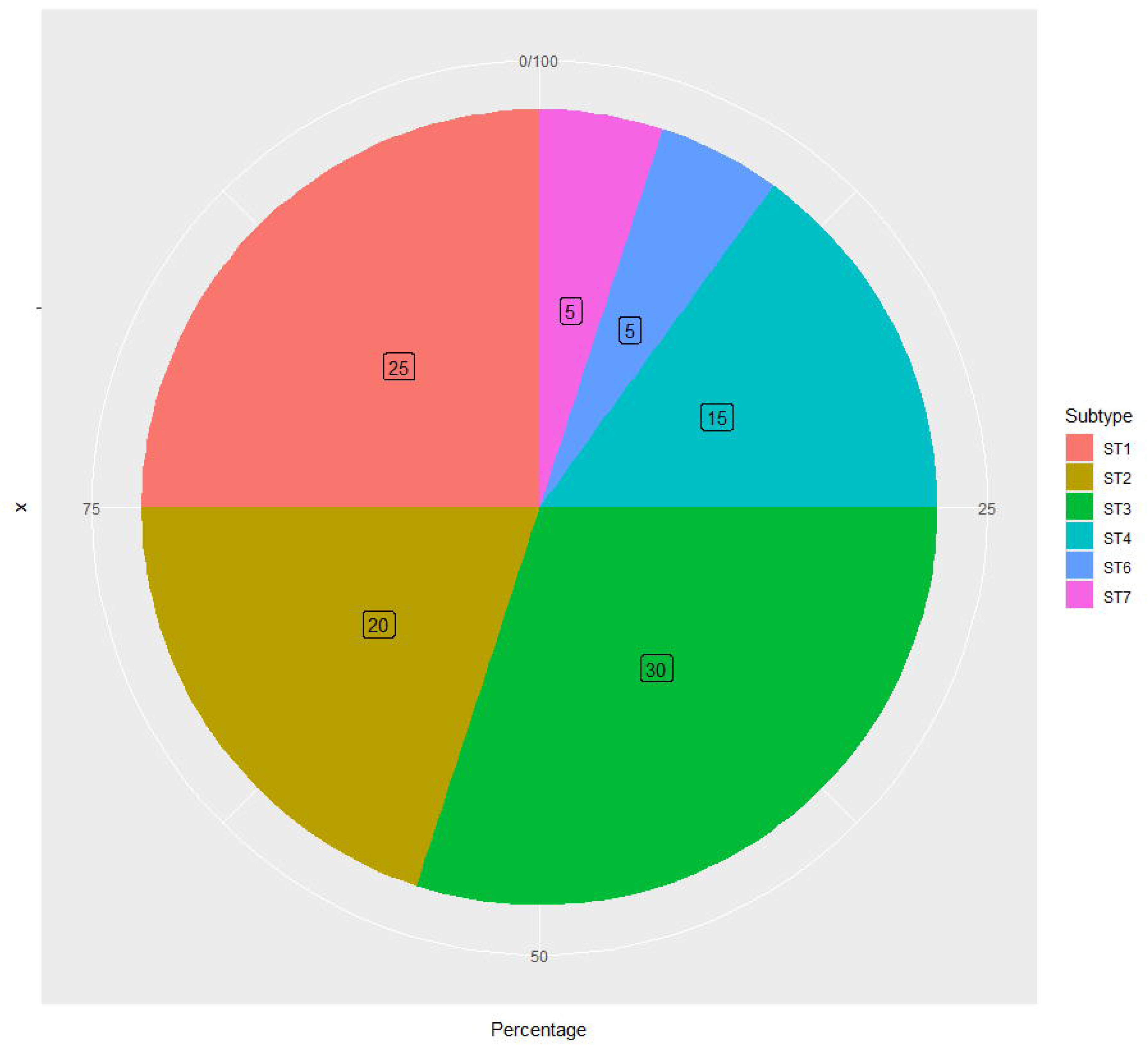
pie chart showing the Subtype distribution of *Blastocystis* in the sampled South East England participants.

### Population parameters and associations with *Blastocystis* colonisation

A Welch T-test measured association with population parameters, and no statistical significance was detected in relation to age (P < 0.05), gender (P < 0.05), or ownership of pets (P < 0.05). Health regimens and general health factors such as alcohol consumption (P < 0.05), use of probiotics (P < 0.05) and BMI (P < 0.05) also showed no statistically significant relationship with *Blastocystis* colonisation. At the same time, there was no statistically significant relationship between *Blastocystis* presence and IBS or UC or between *Blastocystis* colonisation and frequency of defecation.

### Gut microbiome composition and *Blastocystis* occurrence

The 16s prokaryotic gut microbiome composition of the faecal samples was determined by amplification and sequencing of the V3-V4 region of the *SSU* rRNA gene. To monitor changes in alpha diversity influenced by the colonisation status and subtype presence of *Blastocystis*, four different diversity metrics for alpha diversity were measured. Shannon diversity, Simpson diversity, Chao1 diversity and total observed taxa (true richness). The diversity metric scores were first averaged by the colonisation status of *Blastocystis* in the samples (positive/negative) (**Supplementary Figure 2A**). The average diversity score was higher in *Blastocystis*-positive individuals for all four alpha diversity metrics used. Furthermore, for statistical analysis of the colonisation status-averaged alpha diversity scores, post-hoc tests (ANOVA + TukeyHSD/Kruskal-Wallis + Dunn test) showed P-values >0.05 (0.16, 0.35, 0.13, 0.097) for all four comparisons of the diversity score metrics, indicating that the increase in diversity scores of Blastocystis +ve individuals (when comparing across all samples) is non-significant. When *Blastocystis* colonisation was compared across exclusively samples from participants with neither IBS or UC (**supplementary figure 2B**), the average diversity score of Blastocystis +ve samples was either similar (Simpson score) or lower (Shannon, Chao1 and Observed taxa). However, statistical analysis revealed that these changes were all non-significant (p-values: 0.94, 0.74, 0.85, 0.81). Comparing averaged diversity scores for samples +ve for IBS by Blastocystis colonisation (**supplementary figure 2C**) status revealed a clear increase in diversity scores in Blastocystis +ve samples, with statistical analysis revealing that the increase in Chao1 diversity score was significant (p-value 0.032), other metrics were >0.05 (0.14, 0.29, 0.05). Comparing averaged diversity scores for samples +ve for UC by Blastocystis colonisation (**supplementary figure 2D**) showed a similar pattern with samples +ve for Blastocystis having a higher average score across all diversity metrics, with statistical analysis showing these increases were non-significant (p-value: 0.61, 0.38, 0.3, 0.48).

Alpha diversity was then compared across *Blastocystis* subtypes (ST1-ST4)(**figure 2**) using the same four alpha diversity metrics mentioned above. Subtypes 6 and 7 were not included in the analysis as they were detected only in a single instance; thus, no average diversity score could be calculated. Across all four diversity metrics analysed, subtypes 1, 2 and 4 had a higher average diversity score than the average score for samples -ve for Blastocystis colonisation. However, samples +ve for *Blastocystis* ST3 showed an averaged diversity score lower than the negative samples across all four diversity metrics. As subtypes 1,2, and 4 all showed higher averaged diversity scores across all four metrics, a correlation between these subtypes and higher alpha diversity could be implied. However, statistical analysis indicated that these differences were not significant as ANOVA/Kruskal-Wallis comparisons between these groups all showed p-values >0.05 (0.19, 0.28, 0.16, 0.21).

**Figure 2.**
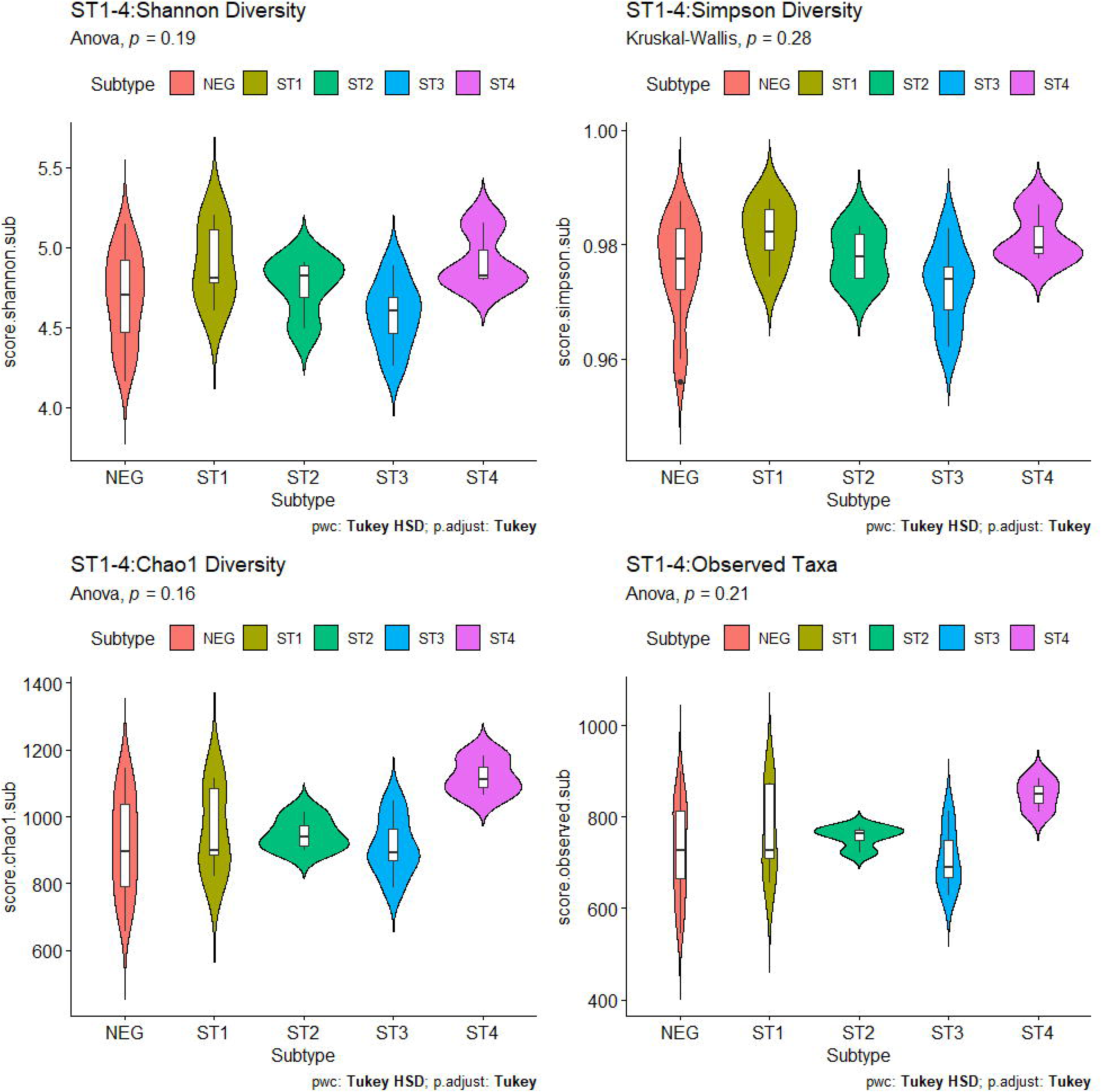
**A**lpha diversity statistical comparisons of diversity metrics, comparisons made between the differing gut microbial content of samples grouped by colonisation between different *Blastocystis* subtypes. Colours are used to denote the four *Blastocystis* subtypes detected (more than once). Diversity metrics used were top-left). Shannon diversity score (top-right). Simpson score (lower-left). Chao1 Diversity, lower-right). Observed taxa/true richness. Statistical analysis subtype groups were performed using ANOVA (normally distributed samples), or Kruskal-Wallis (non-normally distributed samples). Pairwise comparison was performed using the Tukey-HSD test (normally distributed samples) or Dunn Test (non-normally distributed samples). P-values are displayed on their respective plots, P-value >0.05 indicates no significance.

To further compare the differences occurring between the different *Blastocystis* colonisation states observed, principal coordinate analysis (PCoA) ordination methods were used (**Figure 3**) as they allow visual comparisons between the samples based on the complete data within each sample. The Bray-Curtis dissimilarity matrix at the genus level aggregated taxa data of each sample/individual were plotted showing comparisons between Blastocystis colonisation status (**figure 3 left**) and Blastocystis subtype status (**figure 3 right**), colour was used to indicated the colonisation/subtype status and shape was used to indicate the health status of the samples (IBS +ve, UC_IBS+ve or neither condition (healthy)). Statistical comparison of the changes in microbial content between the analysed groups was done using PERMANOVA to test for significance between the positions of the ‘centrons’ of each set of sample metadata. No significant differences between *Blastocystis* +ve and *Blastocystis* -ve participants were detected [PERMANOVA p value 0.211(**figure 3 left**)]. When grouped by subtype metadata (figure 3, right), there was also no significant difference (PERMANOVA P-value 0.348). Nevertheless, when samples were instead grouped by health status (**supplementary figure 6**) statistical analysis revealed that there were statistically significant differences, with a PERMANOVA P-value of 0.008 between the healthy, IBS+ve and UC_IBS+ve groups. The data indicates that IBS and UC/health status are more significant in explaining differences observed in the Kent cohort microbiome data than *Blastocystis* colonisation.

**Figure 3.**
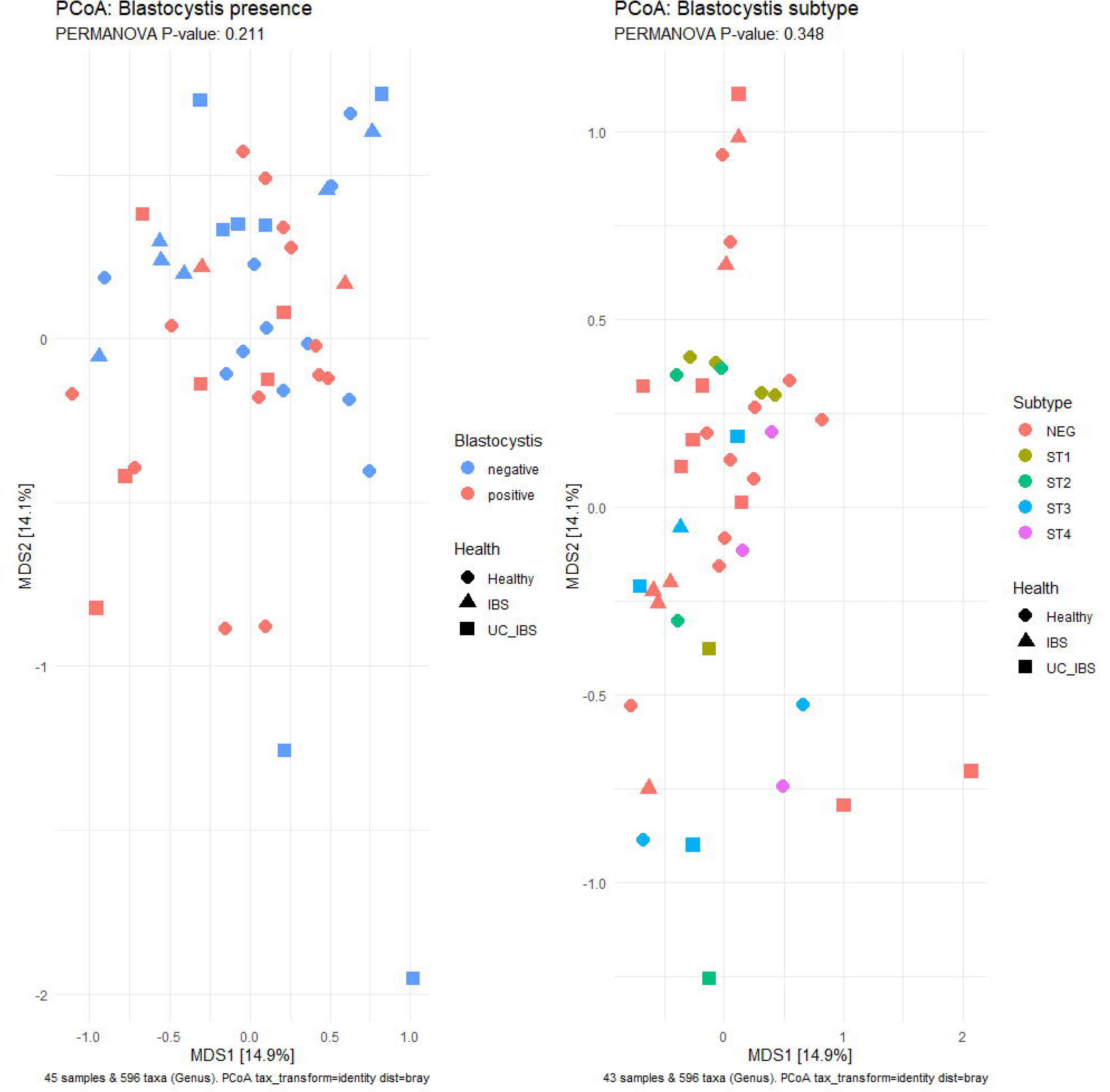
Principle coordinate analysis (PCoA) plot, comparing the microbiome content of samples dependent on *Blastocystis* colonisation or subtype. Taxa abundances/presence are converted into a Bray-Curtis dissimilarity matrix, and the Bray-Curtis positions/ordination scores of each sample are shown in the plot. Samples are grouped and coloured by Left). Colonisation of *Blastocystis* (+ve/-ve) and Right). Colonisation by *Blastocystis* subtype (1-4, (Subtypes which appeared more than once in the dataset. PERMANOVA is used to confirm significance in variation of group centron positions, P-values <0.05 indicate significance.

Compositional plots containing all taxa whose average read count made up more than 1% in each sample, showing the most abundant taxa within each respective sample, with samples grouped by either *Blastocystis* colonisation and *Blastocystis* subtype (**figure 4**), with genus (**figure 4 left column**) and family (**figure 4 right column**) level taxonomic aggregations shown. Samples grouped by *Blastocystis* colonisation at the family level show little variation (**Figure 4 top right**), with a notable increase in the family Bacteroidaceae and reduction in the family Coriobacteriaceae. At the genus level (**figure 4 top left**), there is an increase in *Phocaeicola*_A and simultaneous reduction in *Agathobacter* in the *Blastocystis* +ve group.

**Figure 4.**
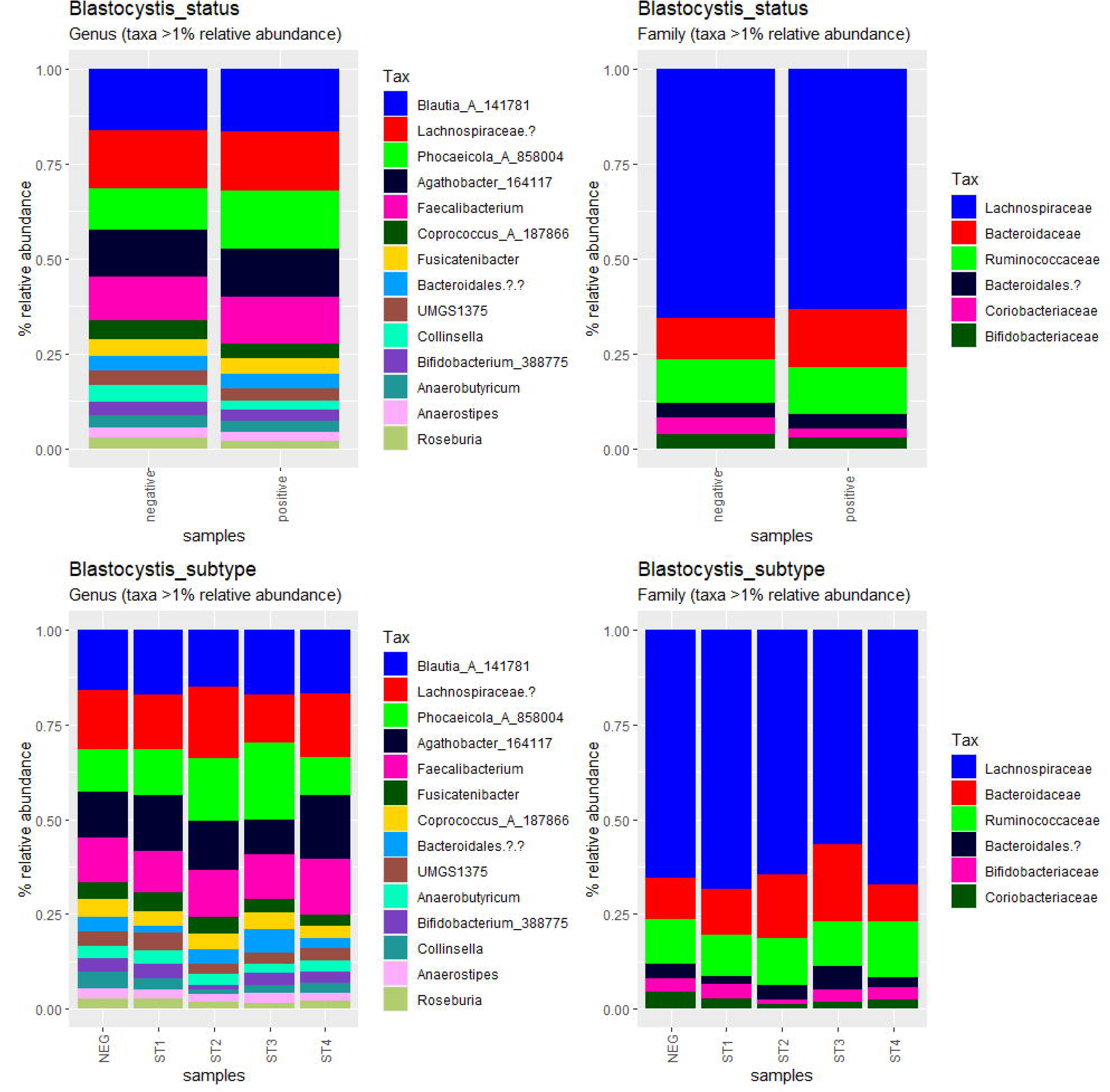
compositional plots showing changes in the relative abundance of taxa which made up >1% total read count of each respective sample. The relative abundance of these >1% taxa are shown in stacked bar plots. Taxa are shown aggregated to Genus (left column) and Family (right column) taxonomic levels. Samples are averaged by Blastocystis colonisation status (positive/negative)(top row) and by *Blastocystis* subtype (subtypes 1-4 and negative) (lower row).

When samples are grouped by *Blastocystis* subtype, more variation is observed. At the family level (**figure 4, lower right**), there is little change observed between the *Blastocystis* subtypes and the negative group, with subtypes ST2 and 3 showing a considerable increase in the Bacteroidaceae family (likely explaining the same change observed in the colonisation status plot (**figure 4, top right)**). At the genus level (**figure 4, lower left**), subtypes 2 and 3 show an increased presence of *Phocaeicola*_A, as well as a slight reduction in *Agathobacter* (the same pattern observed in the colonisation status plots), and all *Blastocystis* subtype groups showed a lowered relative abundance of the genus Collinsella. The simultaneous increase of *Phocaeicola*_A and loss of Agathobacter seems to be a trend strongly observed in samples colonised by Blastocystis subtypes 2 and 3, the most abundant subtypes observed.

#### Metabolome analysis and parasite colonisation

Changes in the metabolome between differing Health statuses (*Blastocystis* colonisation, IBS status and UC status) were first analysed using Principle Component Analysis (PCA) (**figure 5, upper row**). The changes in metabolites based on disease/*Blastocystis* status were monitored using PERMANOVA. *Blastocystis* colonisation showed no significant changes in the metabolites present, with a p-value of 0.118. Changes in metabolite composition between samples grouped by IBS status were also found to be non-significant (p-value 0.051); however, this difference was close to significance, implying a substantial change in metabolite composition. Comparison between samples based on UC status revealed that this was the only status with a significant change observed (p-value 0.018).

**Figure 5.**
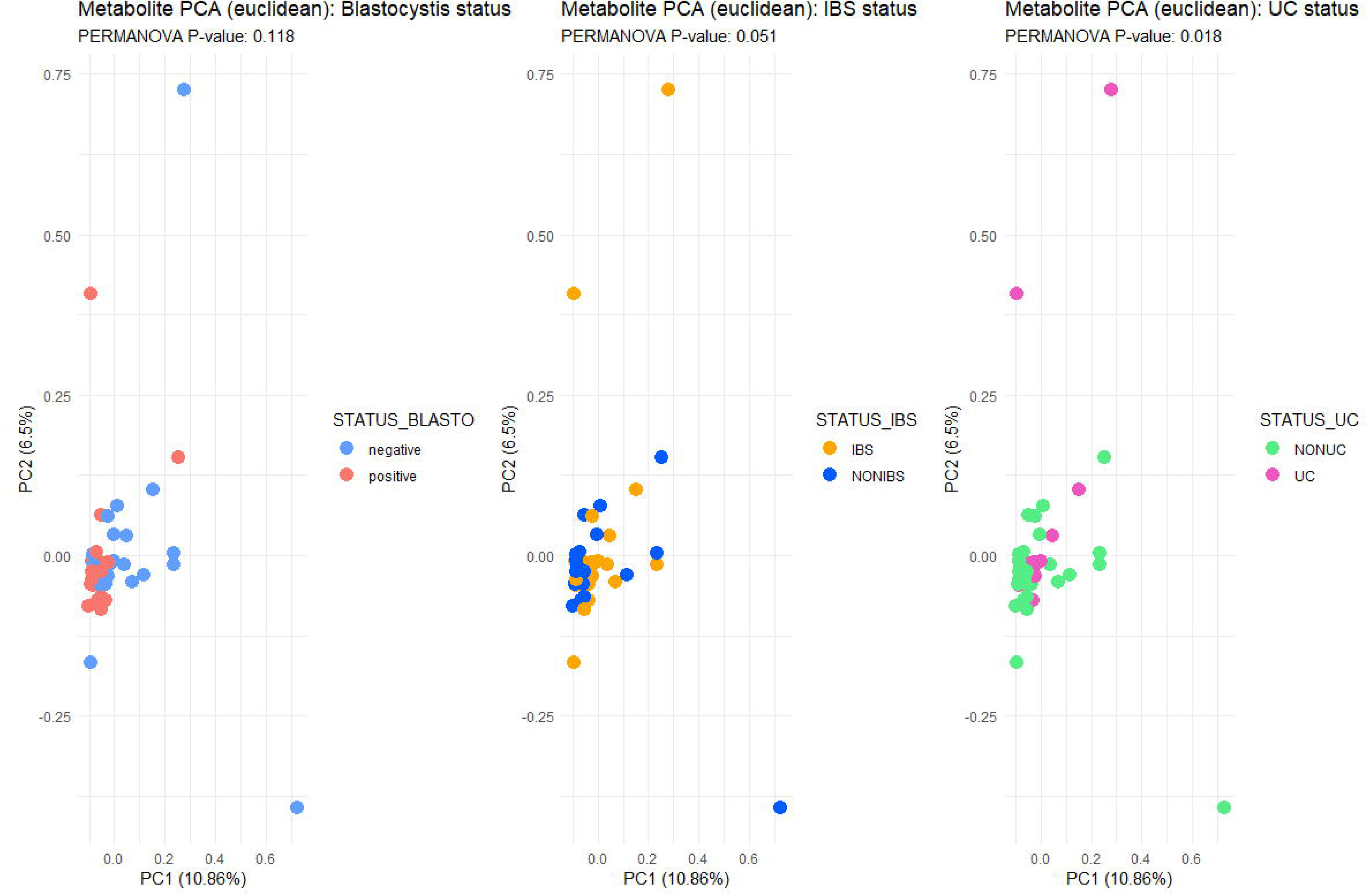
Differences in gut metabolome composition between *Blastocystis* +/-ve samples (left.), Ulcerative Colitis +/-ve samples (centre) and Irritable bowel syndrome +/-ve samples (right). Principle component analysis (PCA) plots (top row) show changes in the overall metabolite composition between samples dependant on their disease/colonisation status (*Blastocystis*, IBS or UC). PERMANOVA was then used to statistically compare the differences in centron position of the two groups in each PCA plot, PERAMOVA p-values of <0.05 show a statistically significant difference in centron position. Differentially expressed metabolites (DEM) are also shown, with log_2_-fold changes in metabolite composition displayed as volcano plots (lower row). The -log_10_ P-value of each metabolite is shown on the y-axis. Labelled and coloured metabolites were those shown to have a statistically significant change in their concentration when comparing between samples grouped by disease/colonisation status (Blastocystis, IBS, UC). Significantly differentially expressed metabolites are coloured based on whether they were found to be upregulated or downregulated in samples positive for the specified disease/colonisation status (*Blastocystis*: red/blue, IBS: orange/blue, UC: pink/green).

Further investigation into the changing occurring in the metabolite composition of samples due to health status/*Blastocystis* colonisation was done using log fold change analysis/volcano plotting. Shown in **figure 5 (lower row)** are volcano plots showing the log_2_ fold change of metabolites, with named and coloured metabolites indicating those who had a statistically significant (significant -log_10_ P value) change in abundance (log_2_ fold change), with colours used to indicate whether the metabolite was upregulated or downregulated in samples positive for Blastocystis/IBS/UC. Analysis of changes in metabolite content considering *Blastocystis* colonisation, ten metabolites had significant changes in their abundance. Adenine, Ethanol and Creatinine were all upregulated. At the same time, Ribose, Ferulate, Acetaminophen, 3-methyl-2-oxovalerate, Thymol, N-Acetylglutamine and Leucine were all downregulated in samples positive for Blastocystis and the opposite true for negative samples. Changes in metabolite abundance based on IBS status revealed that 20 metabolites underwent significant changes in abundance. Leucine, Glucarate, 3-Hydroxyphenylacetate, 6-Hydroxynicotinate, dTTP, Nicotinate, Niacinamide, Kynurenate, 2-Hydroxyisocaproate, N-Phenylacetylphenylalanine and Caffeine were found to be upregulated. Nine metabolites were significantly downregulated; Kynurenine, 2-Hydroxyisovalerate, O-Phosphocoline, Arabinose, 3-Phenyllactate, 1 3 1,3-Dihydroxyacetone, Hydroxyacetone, Proline and Galactonate were all significantly downregulated. Analysis of UC status-associated metabolites showed 14 metabolites significantly changed, only 3-Phenyllactate and Glucose-6-Phosphate were significantly downregulated, while Alanine, Cellobiose, Gentisate, 5-Hydroxytryptophan, 4-Carboxyglutamate, Xanthurenate, Riboflavin, Anserine, 2-Aminoadipate, S-Sulfocysteine, Butyrate and Oxypurinol were upregulated in UC-positive samples. It should be noted that after Benjamini-Hochberg P-adjust these changes were found to be non-significant. All volcano plots underwent p-adjust indicating a potential of false positives in these plots.

### Microbiome and metabolome comparative analysis and *Blastocystis* colonisation

To compare the changes observed in the microbiome and metabolome, a combination of Linear discriminant analysis and Spearman’s rank correlation analysis was performed (figure 6). The significantly discriminant taxa were then compared with the metabolites which had a statistically significant log fold change (as determined in figure 5), the potential correlation between linearly discriminant ‘biomarker’ taxa and the health/Blastocystis status associated metabolites, was evaluated using spearman’s rank correlation. As shown in the LEfSe plot in **figure 6A**, multiple taxa were linearly discriminant for either value of Blastocystis status (Positve/negative), these taxa exceeded the significance threshold for both *Blastocystis* +ve and -ve colonisation status sample groups. Six taxa were significantly discriminant for the *Blastocystis* -ve status, with the three highest LDA scores for the genera Collinsella, UMGS1375 (of the family *Lachnospiraceae*) and *Faecalibacillus*. Fifteen taxa were significant discriminants for the *Blastocystis* +ve status. The three highest LDA scores were for and unclassified member of the family Pseudomonadaceae, the genera Dysombacter and an unclassified member of the family Oscillospiraceae. These significantly discriminant taxa were correlated with the significant metabolite data. The strongest correlations between the Blastocystis -ve discriminant taxa were a strong positive correlation between the genera Collinsella and the amino acid Adenine, and between the genera Klenkia and the amino acid Leucine. The strongest correlations between Blastocystis +ve discriminant taxa were a strong negative correlation between the unclassified member of the family CAG-552 (member of the order Clostridiales) and the amino acid Leucine, and a strong negative correlation between the genera Enterocloster and both the amino acid Leucine and Ferulate/Ferulic acid.

**Figure 6.**
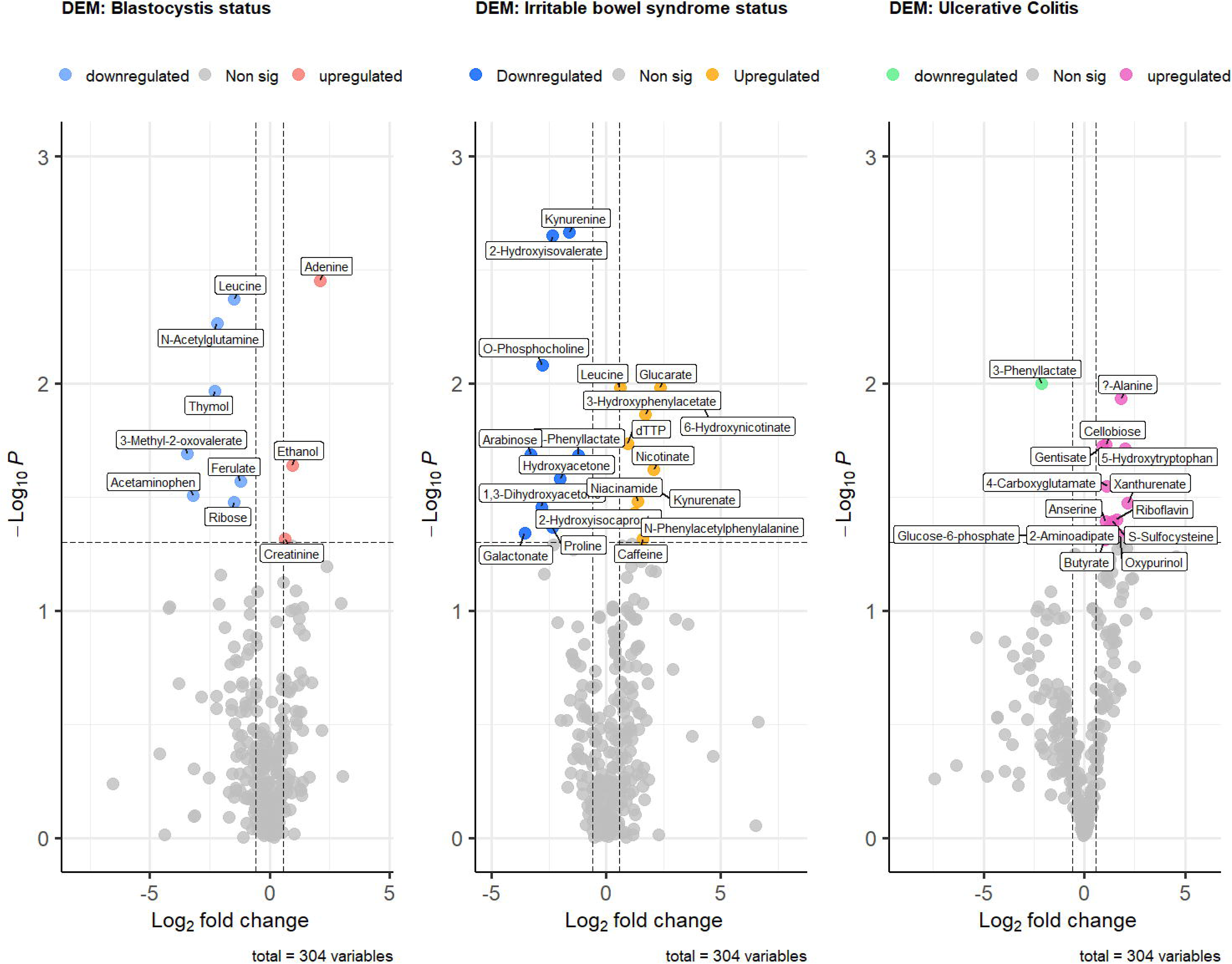
Cross-examination of the 16s microbiome sequencing data (aggregated to genus level) and metabolome data. Linear discriminant analysis Effect Size (LEfSe) plots showing the taxa with significant linear discriminant effect size scores (LDA score/x-axis)(any LDA score >2/<-2 is significant, although in UC and IBS plots the minimum LDA threshold was increased to and LDA score of 3). Spearman’s rank correlation plots show the correlation of the significantly discriminant taxa with metabolites which were identified (via volcano plot) to be significantly differentially expressed dependant on disease/colonisation status of the samples (significant changes in their concentration). The LEfSe plots are coloured to show whether each taxa was a discriminant taxa for either the +ve or -ve status for each disease/colonisation status (Blastocystis, IBS, UC). Spearmans rank correlations were also coloured to indicate their relationship with the disease/colonisation status, with the colour indicating whether their was a positive or negative correlation between the taxa and the metabolite. Colour coordination. Blastocystis status (LEfSe = red: Blastocystis +ve, blue: Blastocystis -ve, Spearmans = red: positive correlation, blue: negative correlation), irritable bowel syndrome (IBS) status (LEfSe = orange: IBS +ve, blue: IBS -ve, Spearmans = Orange: positive correlation), Ulcerative Colitis (UC) status (LEfSe = pink: UC +ve, Green: UC -ve, Spearmans = pink: positive correlation, green: negative correlation).

Thirty-two taxa were found to be significantly discriminant for IBS-ve samples, with the three strongest discriminant taxa (highest LDA score) being the *genera* Clostridium_T, CAG-273 (of the class Clostridia) and ER4 (also known as *Oscillibacter*). A wide range of metabolites had strong correlations with the IBS-ve discriminating taxa, with both ER4 and SFLA01 (of the order Oscillospirales) having strong positive and negative correlations with 6 differentially expressed metabolites. The metabolites which had correaltions with the widest range of IBS-ve discriminant taxa were 1,3-Dihydroxyacetone (a saccharide), the amino acid Proline and Kynurenine (a metabolite of the amino acid Tryptophan), these metabolites all had strong negative correlations with multiple the IBS -ve discriminant taxa. On the other hand, Nicotinate (a vitamin B3 derivative), the amino acid Leucine and 2-Hydroxyisocaproate (a metabolite resulting from Leucine metabolism) had strong positive correlations with multiple taxa discriminant for the IBS-negative status; therefore, these metabolites are likely strongly correlated with samples negative for IBS. Taxa significantly discriminant for IBS+ve samples were fewer in number with only nine taxa, an unclassified member of the family Anaerotignaceae, the genera Massilia and Curtobacterium had the highest LDA scores and were the strongest discriminant taxa for IBS+ve samples. As expected the strongest metabolite-taxa correlations were the inverse of the IBS-ve correlated metabolites with 1,3-Dihydroxyacetone and Galactonate (negatively correlated to IBS-ve taxa) found positive correlations with IBS+ve taxa. The same was true for Nicotinate, Leucine and 2-Hydroxyisocaproate, which were all negatively correlated with IBS+ve taxa.

Analysis of taxa associated with UC status found twenty-five taxa significantly discriminant for the UC-ve status, with the strongest discriminant taxa being an unclassified member of the family Lachnospiraceae, Clostridium_T and Mediterranebacter. A smaller number of metabolites were strongly correlated with the UC-ve discrimiant taxa, Anserine, Butyrate (a short chain fatty acid), Riboflavin (vitamin B2), S-Sulfocysteine and the amino acid Alanine being all negatively correlated with the UC -ve discriminant taxa. Only 3-Phenyllactate (Phenylacetic acid) was strongly positively correlated with the UC -ve discriminant taxa. Seventeen taxa were found to be significantly discriminant for the UC +ve status, with the strongest discriminant taxa being unclassified members of the families Ruminococcaceae and Anaerotignaceae, and the genera Massilia. Multiple metabolites were found to be strongly correlated with the UC +ve discriminant taxa, notably a strong negative correlation between Glucose-6-phosphate and multiple UC +ve discriminant taxa, and a very strong positive correlation between almost every UC +ve discriminant taxa and the metabolite S-Sulfocysteine, which was negatively correlated with multiple UC -ve discriminant taxa.

## Discussion

This pilot study represents the first integrated microbiome–metabolome analysis of *Blastocystis* colonisation in South East England, assessing its prevalence, subtype distribution, and associations with gut microbial and metabolic profiles in individuals with and without IBS or UC. *Blastocystis* was detected in 47.8% of the cohort, with ST3 the most common subtype, consistent with reports from other UK and European populations. No significant associations were observed between *Blastocystis* colonisation and demographic or clinical variables, including IBS or UC, which aligns with previous studies in high-income settings where exposure risk factors are more homogenous.

Alpha diversity was consistently higher in *Blastocystis*-positive individuals across all diversity metrics, with statistical significance reached in the IBS subgroup for Chao1 richness. Subtype comparisons suggested that ST1, ST2, and ST4 may be associated with greater diversity, while ST3 showed reduced diversity, though these differences were not statistically significant. Beta diversity analysis indicated that *Blastocystis* colonisation had a limited impact on overall microbiota structure, whereas IBS and UC status were substantial drivers of community composition. Taxonomic shifts in *Blastocystis*-positive individuals included increases in *Phocaeicola_A* and reductions in *Agathobacter* and *Collinsella*, consistent with previous associations of *Blastocystis* with more stable, health-associated microbiomes (BICLOT, 2023).

Metabolomic profiling showed no significant global differences in metabolite composition between *Blastocystis*-positive and -negative individuals. However, exploratory volcano plot analysis identified ten metabolites with unadjusted differential abundance, including lower levels of leucine and kynurenine in *Blastocystis*-colonised individuals (Figure 5). These trends were further explored through taxon–metabolite correlation analysis.

One of the most notable findings was the negative correlation between *Blastocystis*-associated taxa and leucine, as well as its microbial metabolite 2-hydroxyisocaproate. Leucine is a branched-chain amino acid involved in gut barrier maintenance, immune regulation, and microbial cross-feeding. Interestingly, in this study, leucine was also positively correlated with IBS status, suggesting that elevated levels may reflect, or contribute to, disease-associated dysbiosis. Although leucine is often considered beneficial and is used in supplements aimed at improving gut function, its metabolism by gut bacteria can lead to the accumulation of pro-inflammatory intermediates. Prior studies have shown that microbial dysregulation of branched-chain amino acid pathways, including leucine, can disrupt gut homeostasis and immune signalling. Therefore, the observed inverse relationship between *Blastocystis*-linked taxa and leucine abundance may reflect a shift towards a microbial community that either produces less leucine or promotes its more efficient utilisation, potentially reducing inflammatory burden (Cruz, Oliveira and Gomes-Marcondes, 2017). These findings align with the broader concept that *Blastocystis* colonisation may support or reflect microbial configurations associated with metabolic balance.

Equally important was the observed negative correlation between *Blastocystis***-**associated taxa and kynurenine, a metabolite of the tryptophan pathway with well-established roles in immune regulation and inflammation. The kynurenine pathway leads to the production of NAD+ and several immunomodulatory intermediates, and its dysregulation has been implicated in conditions ranging from metabolic liver disease to depression and cancer. Recent studies have demonstrated that gut-derived kynurenine is linked to hepatic steatosis and fibrosis in NAFLD, underscoring its significance as both a biomarker and a mediator of systemic disease. The finding that several *Blastocystis*-associated taxa were negatively correlated with kynurenine in this study adds further weight to the growing body of evidence linking *Blastocystis* colonisation to gut environments with lower inflammatory potential (Rojas-Velázquez *et al*., 2022). Although the mechanisms behind this association remain to be elucidated, the data suggest that *Blastocystis* may interact with the microbiota in ways that indirectly suppress kynurenine production or activity.

While these associations are compelling, none of the metabolite differences remained statistically significant after adjustment for multiple testing, and beta diversity analyses confirmed that UC had a more substantial effect on metabolite composition than *Blastocystis* colonisation. Still, the consistent correlations between *Blastocystis*-linked taxa and key metabolites such as leucine and kynurenine suggest potentially meaningful interactions between colonisation status and metabolic function that merit further investigation.

This study is constrained by its small sample size, which limits the ability to detect subtle differences and assess subtype-specific effects in detail. The cross-sectional design prevents any inference of causality or directionality, and the lack of detailed dietary and inflammatory marker data is an additional limitation. Future research should focus on longitudinal designs with repeated sampling and integrated metagenomic, metabolomic, and host immune profiling (Figueiredo *et al*., 2025). Functional studies using cultured isolates and *in vitro* models will be crucial to determine whether *Blastocystis* directly modulates gut metabolite production or influences it by shaping the broader microbial community. Expanding this research to larger, more diverse populations will be vital to better understanding the ecological and clinical significance of this common yet enigmatic protist.

While subtypes such as ST7 have shown pathogenic potential in *in vitro* studies (Deng, Lee and Tan, 2022)(Yason *et al*., 2019), exhibiting mechanisms like increased protease activity, suppression of nitric oxide production, and stimulation of proinflammatory cytokines, conclusive *in vivo* evidence is still lacking (Deng and Tan, 2025). In contrast, other subtypes, notably ST3 and ST4, are frequently detected in asymptomatic individuals, particularly in Europe (Deng *et al*., 2022)(Tito *et al*., 2019).

## Conclusions

This pilot study offers a first look at the prevalence, subtype distribution, and ecological associations of *Blastocystis* in South East England, highlighting its widespread presence and potential links to gut microbial and metabolic profiles. While no major differences in microbiome or metabolome composition were detected between *Blastocystis*-positive and - negative individuals, the observed negative correlations between *Blastocystis*-associated taxa and metabolites such as leucine and kynurenine point to potentially beneficial metabolic environments. Given leucine’s complex role in gut immunity and its positive correlation with IBS in this dataset, and kynurenine’s established involvement in inflammation and systemic disease, these findings raise important questions about whether *Blastocystis* plays a stabilising role in gut metabolic networks. Future studies should focus on larger, longitudinal cohorts incorporating high-resolution metagenomics, targeted metabolomics, dietary data, and inflammatory markers, alongside functional *in vitro* models using cultured isolates. Such approaches will be essential to determine whether *Blastocystis* modulates microbial metabolism directly, reflects underlying gut health, or acts as a passive coloniser with subtype-dependent effects. Clarifying these roles will be critical for understanding whether *Blastocystis* represents a marker, mediator, or modulator of gut ecosystem health.

## Data Availability

All data produced in the present study are available upon reasonable request to the authors

## Acknowledgments

We would like to thank the volunteers for providing the samples. Special thanks to Dr. Raul Tito for his guidance on the analysis and to Przemek Wieckowski for help with metabolite analysis scripts. W.J.S.E. was supported by the South Coast Biosciences Doctoral Training Partnership SoCoBio DTP BBSRC BB/T008768/1 and J.M.N. by a scholarship from Kent Health.

---

**Supplementary Figure 1:** rarefaction curve of samples involved in the study.

**Supplementary Figure 2: A**lpha diversity statistical comparisons of diversity metrics, comparisons made between the differing gut microbial content of samples grouped by *Blastocystis* colonisation in samples with differing health statuses. Overall *Blastocystis* colonisation status regardless of health status (top left), *Blastocystis* colonisation in healthy samples (top right), *Blastocystis* colonisation in IBS +ve samples (lower left) and *Blastocystis* colonisation status in UC+ve samples (lower right). Diversity metrics used were top-left). Shannon diversity score, Simpson score, Chao1 Diversity, Observed taxa/true richness. Statistical analysis of subtype groups was performed using ANOVA (normally distributed samples), or Kruskal-Wallis (non-normally distributed samples). Pairwise comparison was performed using the Tukey-HSD test (normally distributed samples) or Dunn Test (non-normally distributed samples). P-values are displayed on their respective plots, P-value >0.05 indicates no significance.

**Supplementary Figure 4:** compositional plots showing changes in the relative abundance of taxa which made up >1% total read count of each respective sample. The relative abundance of these >1% taxa are shown in stacked bar plots. Taxa are shown aggregated to Genus (top row) and Family (lower row) taxonomic levels. Samples are averaged by *Blastocystis* colonisation status (positive/negative)(top row) and by *Blastocystis* subtype (subtypes 1-4 and negative)(lower row).

**Supplementary Figure 5:** compositional plots showing changes in the relative abundance of taxa which made up >1% total read count of each respective sample. The relative abundance of these >1% taxa are shown in stacked bar plots. Taxa are shown aggregated to Genus (top row) and Family (lower row) taxonomic levels. Samples are not averaged and the complete sample set are grouped by *Blastocystis* colonisation status (positive/negative) (far left column), *Blastocystis* subtype (subtypes 1-4 and negative)(left column), IBS status (positive/negative)(right column), and UC status (positive/negative)(far right column).

**Supplementary Figure 6:** Principle coordinate analysis (PCoA) plot, comparing the microbiome content of samples dependent on sample health status (IBS positive, IBS and UC positive or lack of either (healthy)). Taxa abundances/presence are converted into a Bray-Curtis dissimilarity matrix, and the Bray-Curtis positions/ordination scores of each sample are shown in the plot. Samples are grouped and coloured by health status. PERMANOVA is used to confirm significance in variation of group centron positions, P-values <0.05 indicate significance.

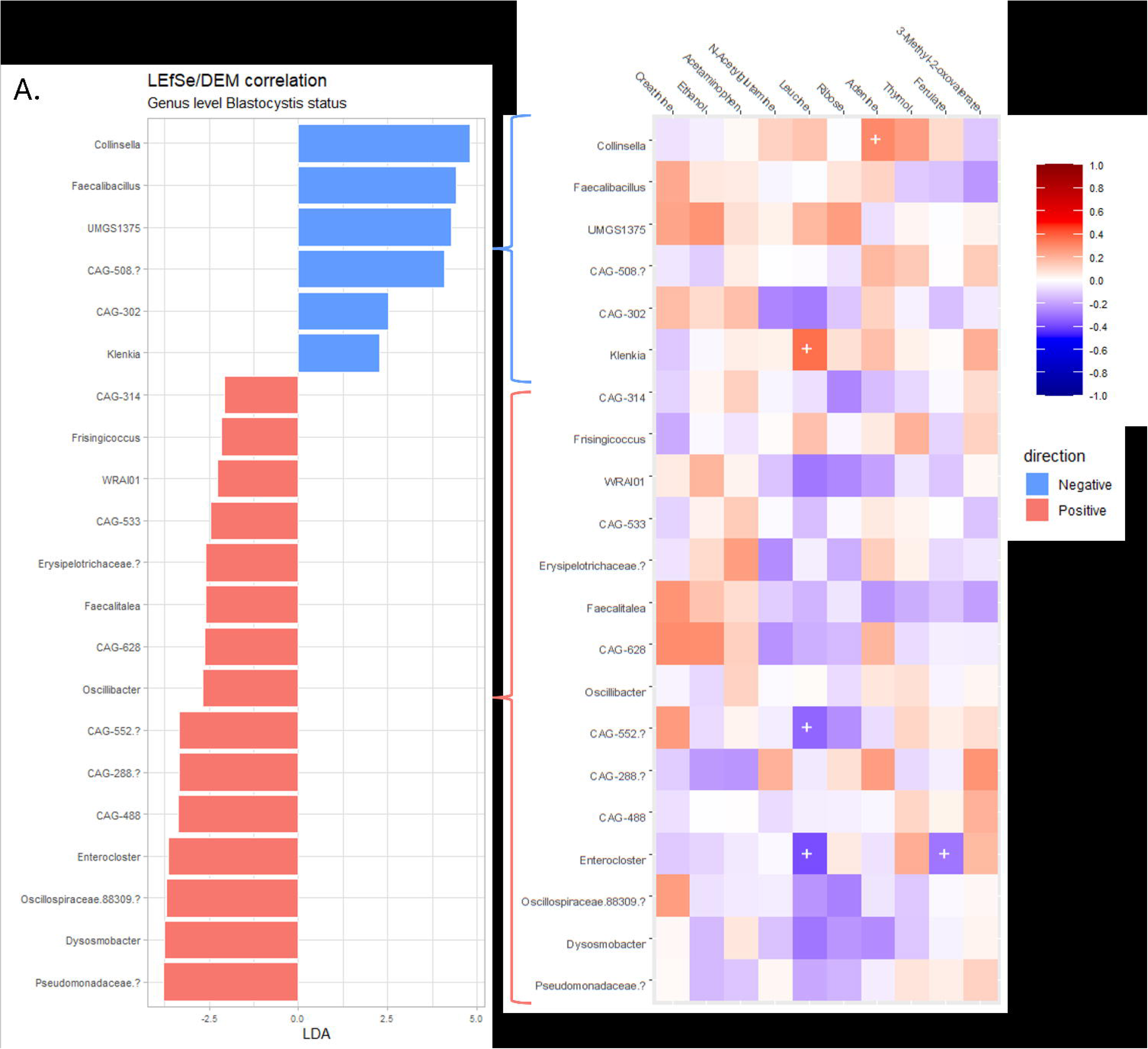
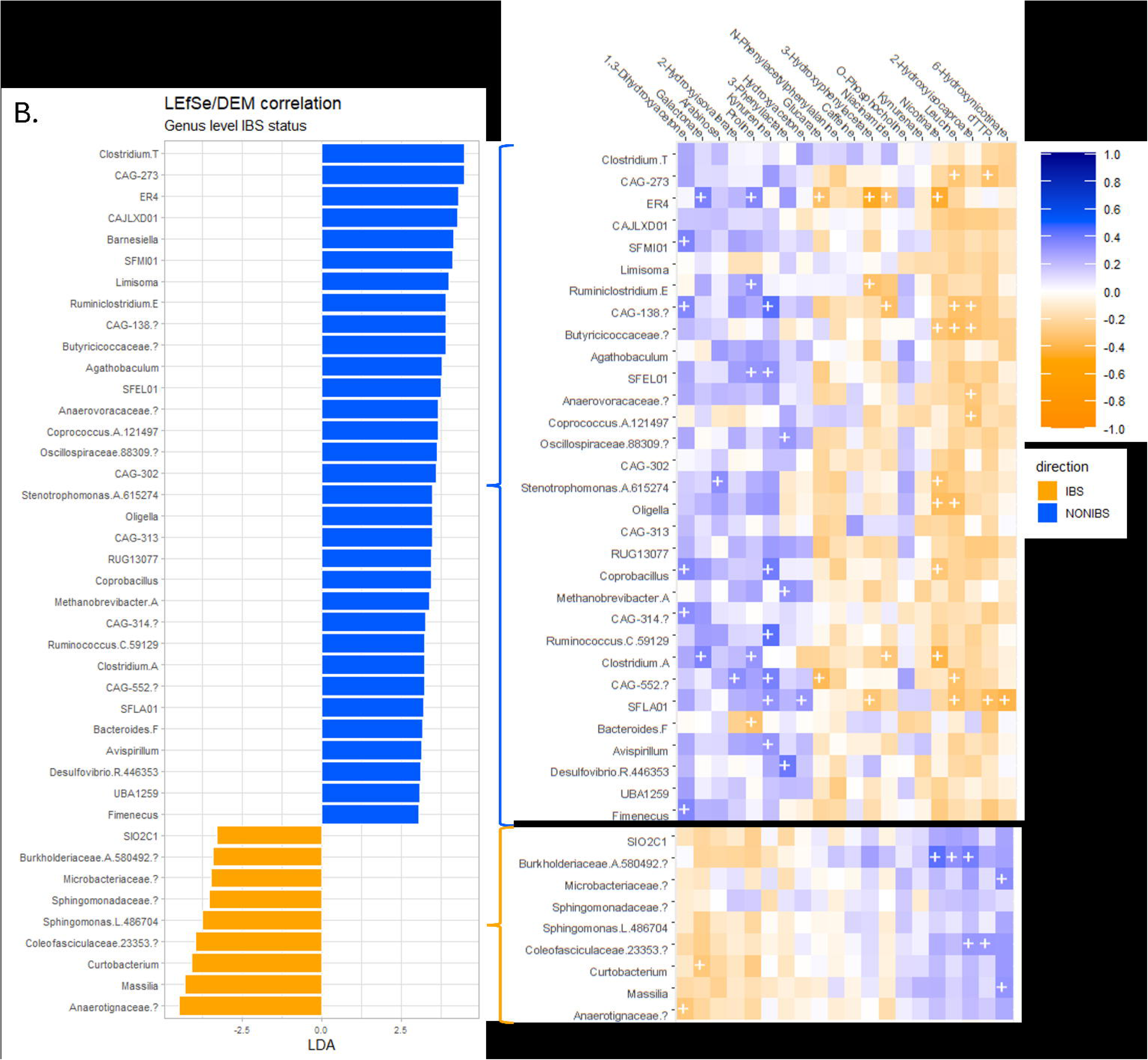
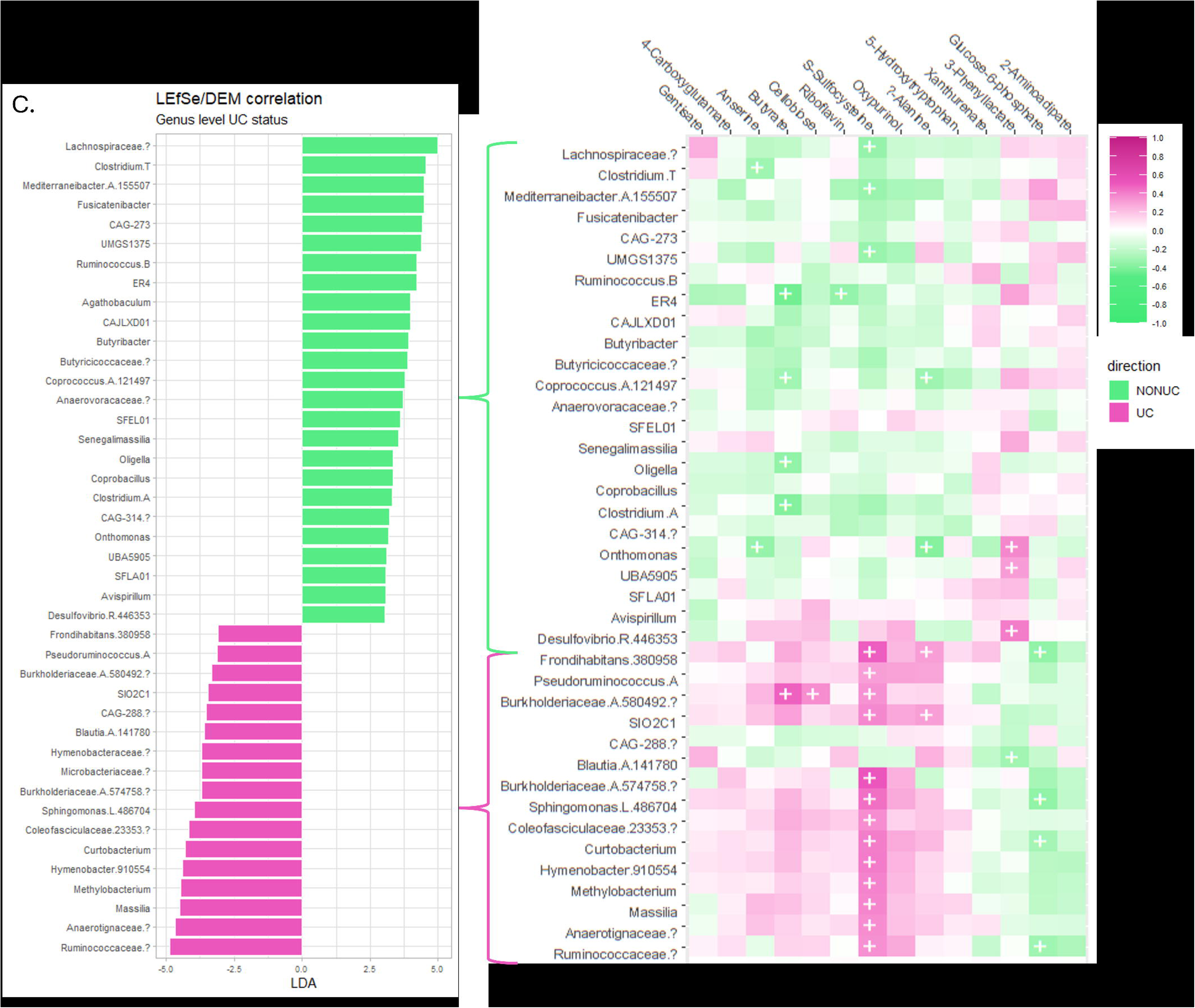

