## Supplementary figures and images for "Exploring *Blastocystis* and Gut Health: A Pilot Investigation of Microbiome and Metabolomic Signatures in a UK Cohort"

### Supplementary Figure 2

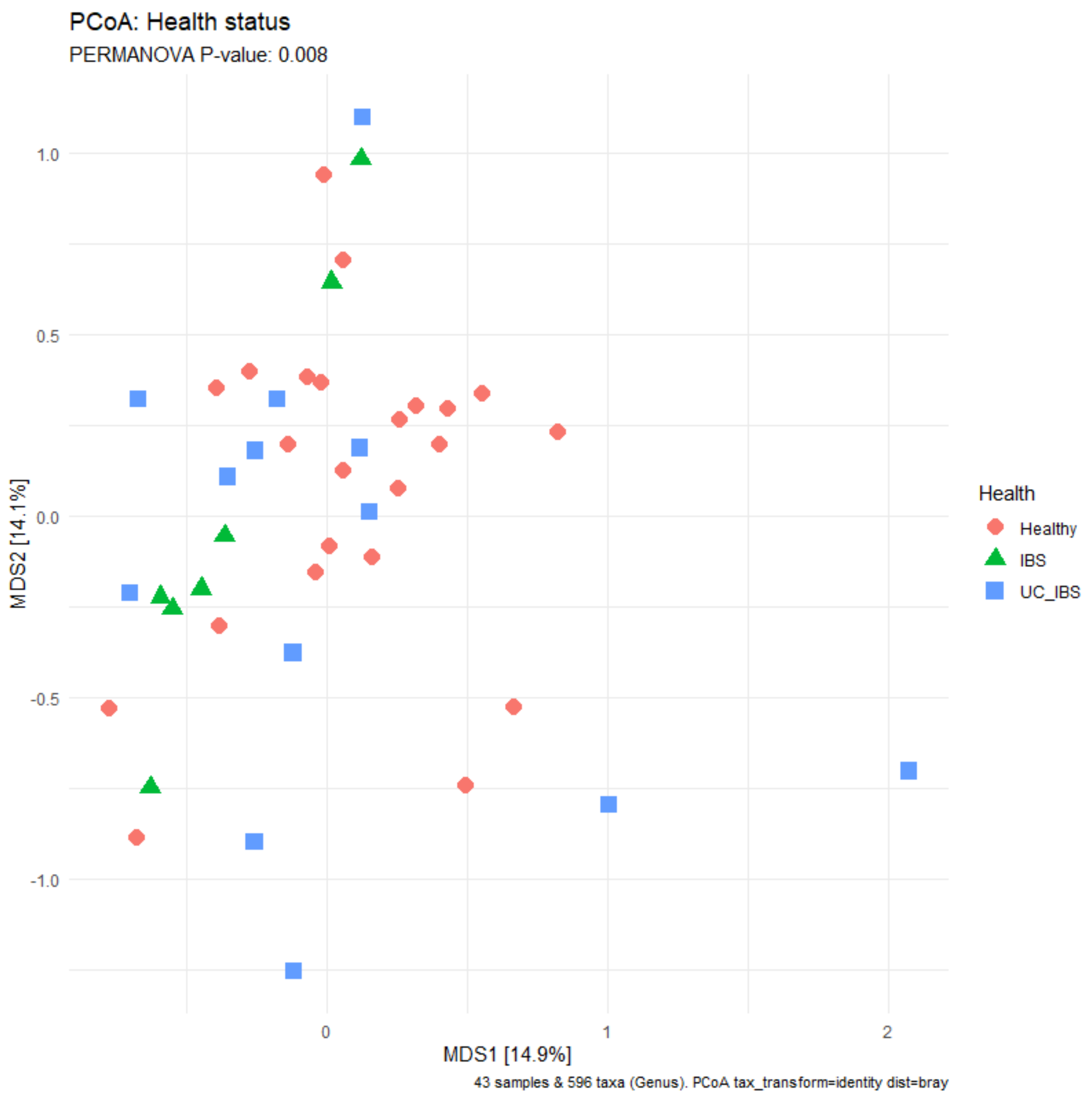

### Supplementary Figure 4

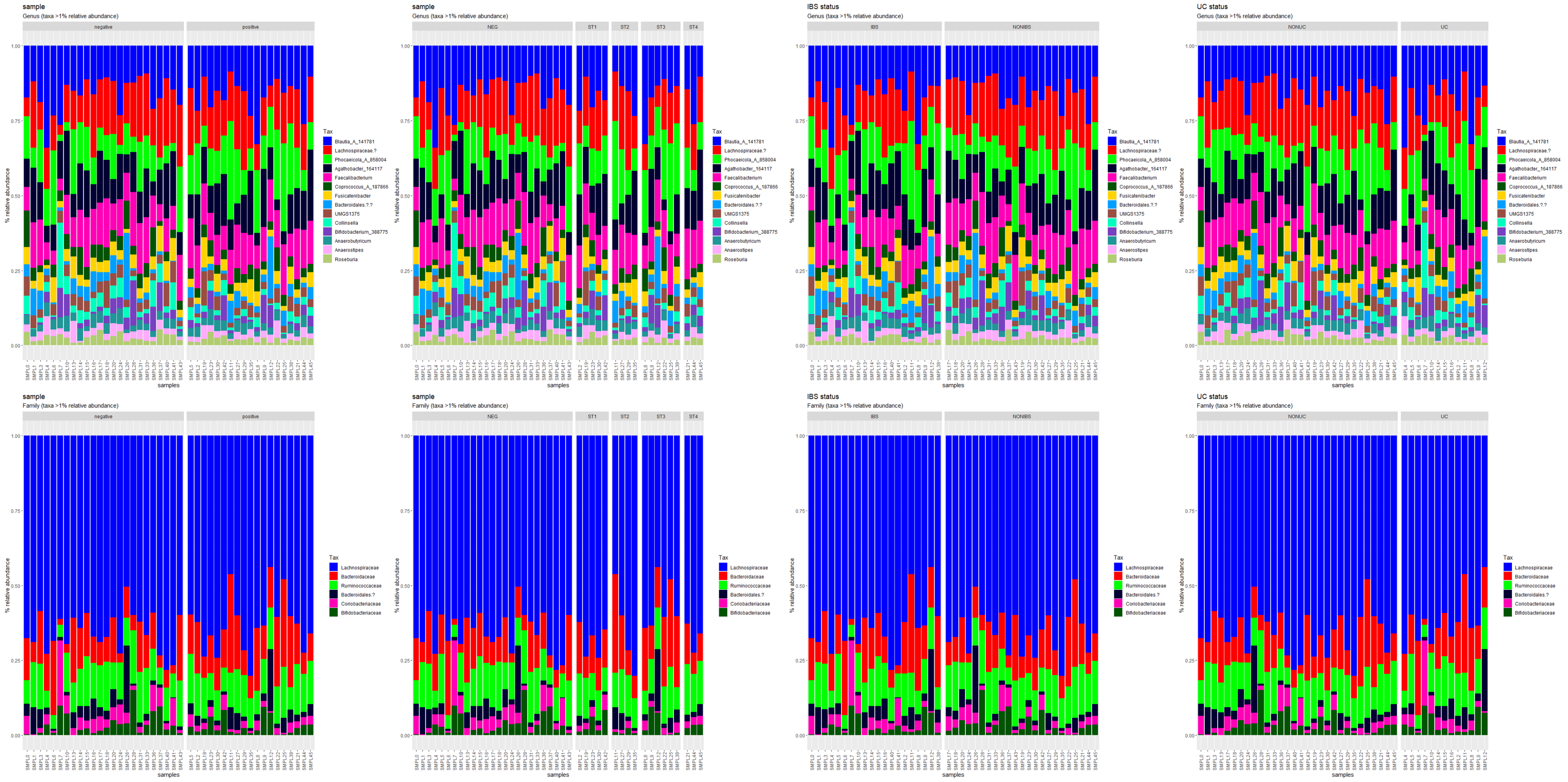

### Supplementary Figure 5

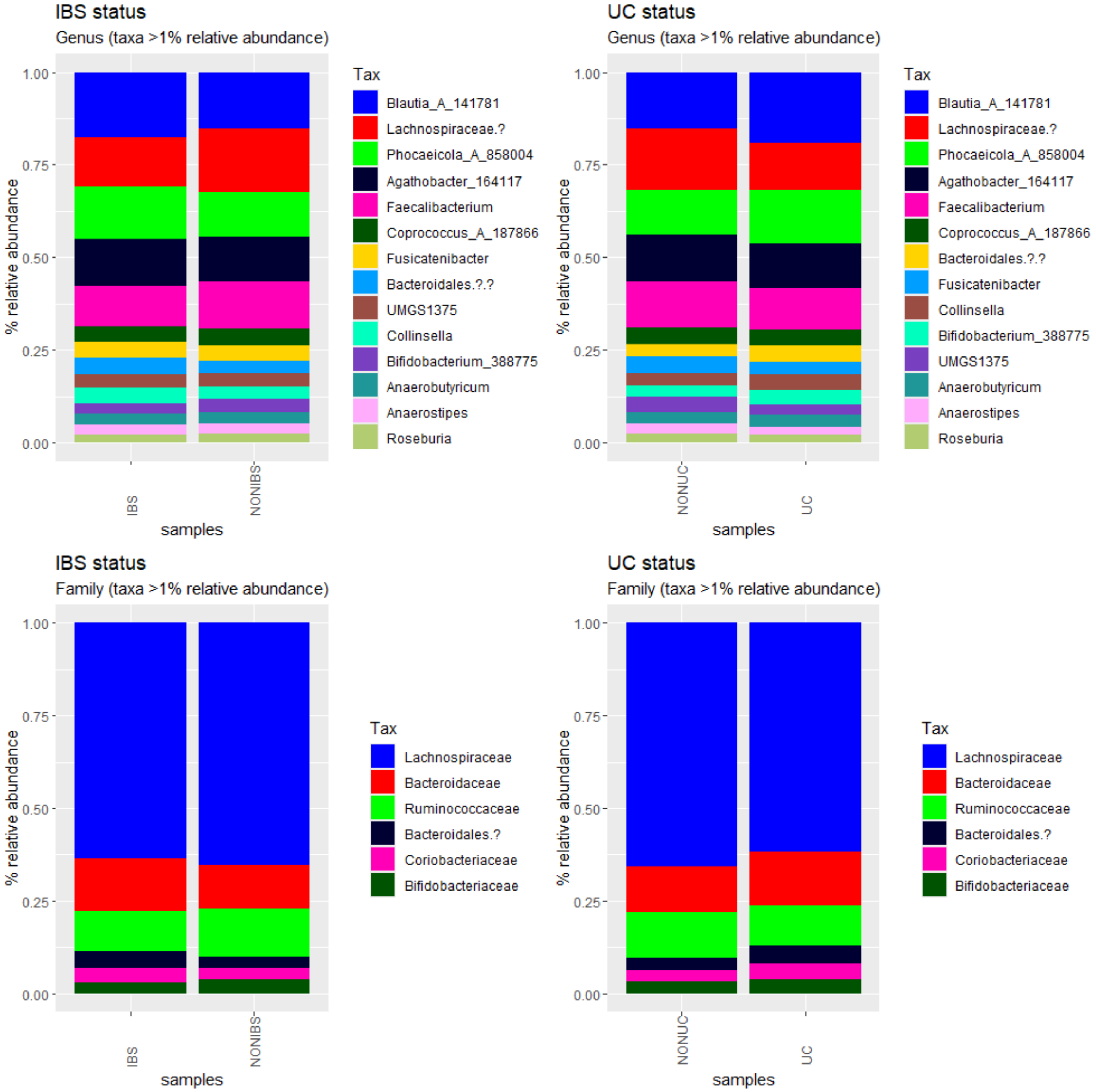

### Supplementary Figure 6

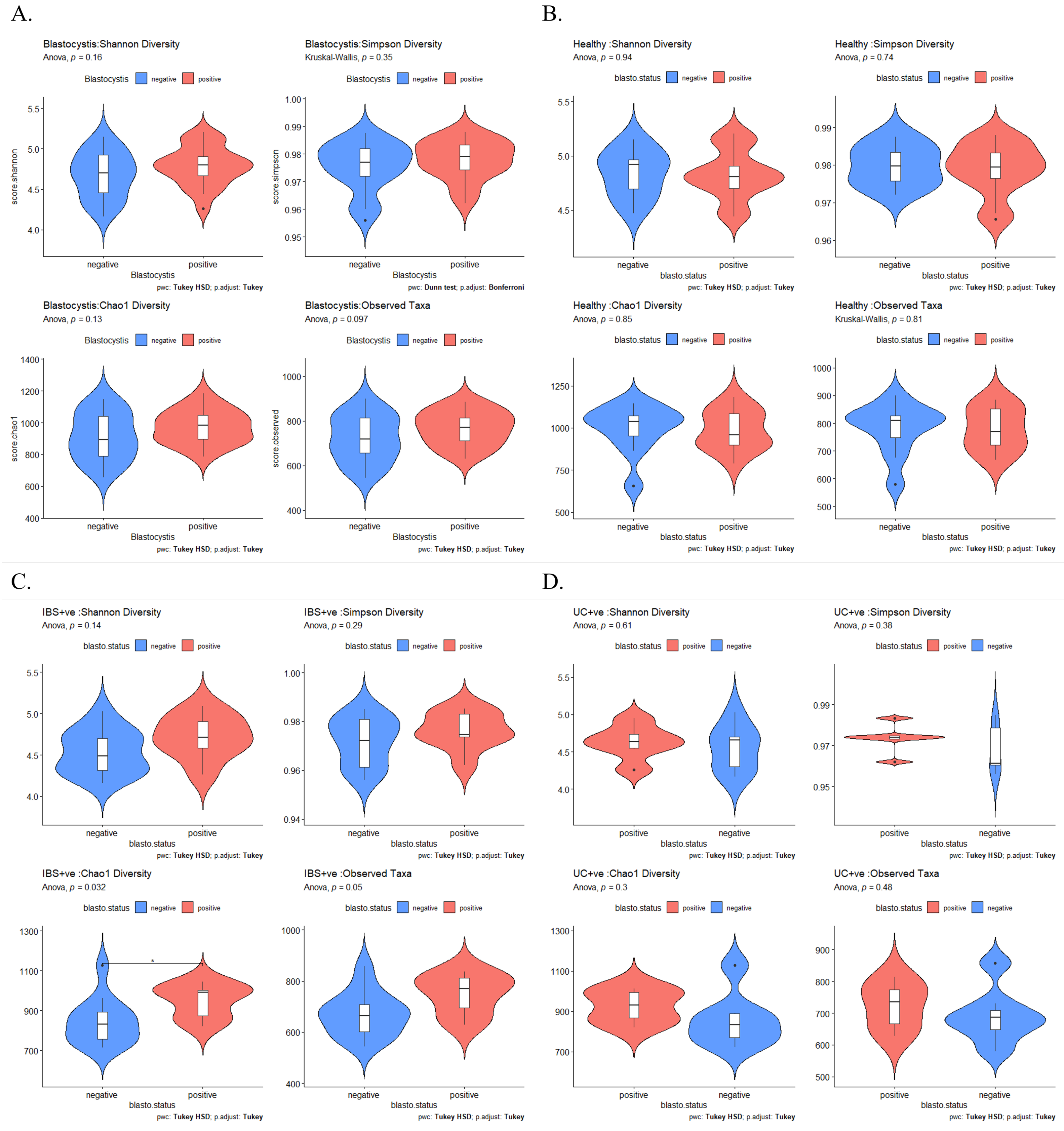
